# Structural and functional signatures of executive deficits after early use of cocaine depend upon route of administration

**DOI:** 10.1101/2020.06.12.20129577

**Authors:** Alethia de la Fuente, Sofía Schurmann Vignaga, Pilar Prado, Rosario Figueras, Lucia Lizaso, Facundo Manes, Marcelo Cetkovich, Enzo Tagliazucchi, Teresa Torralva

**Affiliations:** Buenos Aires Physics Institute (IFIBA) and Physics Department, University of Buenos Aires; Institute of Cognitive and Translational Neuroscience (INCYT), INECO Foundation, Favaloro University, Buenos Aires, Argentina; National Scientific and Technical Research Council, Buenos Aires, Argentina

**Keywords:** Cocaine, Routes of administration, Executive function, Attention, MRI/fMRI

## Abstract

**Background:** There is growing evidence linking cocaine consumption with a broad spectrum of neurocognitive deficits. Despite of evidence suggesting that the route of administration should be taken into account to assess the short and long term effects of cocaine consumption, to our knowledge no study to date has characterized clinically relevant neuropsychological variables along with physiological variables separately in populations of individuals with histories of smoked cocaine dependence (SCD) and insufflated cocaine hydrochloride dependence (ICD).

**Methods:** The present study examined a sample of (a) 25 participants who fulfilled criteria for SCD, (b) 22 participants who fulfilled criteria for ICD, and (c) 25 healthy controls matched by age, gender, education, and socioeconomic status. An exhaustive neuropsychological battery was used to assess different cognitive domains (attention, executive functions, fluid intelligence, memory, language and social cognition). We complemented this neuropsychological assessment with the acquisition and analysis of structural (MRI) and functional (fMRI) neuroimaging data.

**Results:** Different routes of administration led to equally different profiles of neurocognitive impairment, with the SCD group being specifically associated with deficits in attention and executive functions. Consistent with risk models, executive function-attention deficit is better explained for age and age onset of consumption initiation. SCD also presented reduced grey matter density relative to ICD in the bilateral caudate, a key area for executive functions and attention. Connectivity between left caudate and inferior frontal regions mediates performance-structure association.

**Conclusions:** Cocaine routes of administration are associated to a differential profile that may not be due direct effects of stimulant action but also driven by cognitive and biological differences in key executive functioning and attention areas. This point the critical importance of the routes of administration. This information could inform clinical management and should be taken into account in clinical research.

## Introduction

The route of administration of cocaine plays a critical role in the latency, magnitude, and duration of the acute effects, as well as in behavioral variables such as use frequency and intake per session ^1–3^. Faster routes of administration (e.g. smoked cocaine [SC]) present higher abuse potential due to stronger reinforcement compared to routes with slower onset of effects (e.g. insufflated cocaine [IC]) ^4–7^. These differential effects are known to be at least partially mediated by a combination of pharmacokinetic factors and modifications in dopaminergic signaling^8,9^. The neuropsychological and neurophysiological bases of cocaine consumption and abuse remain controversial ^10^, partially because most clinical and preclinical studies to date failed to take into account effects that are specific to different routes of administration ^11–14^. Understanding the relationship between cocaine consumption and the route of administration is critical to assess the effects of different forms of cocaine abuse. For instance, addiction to SC develops earlier and is more difficult to overcome ^15–17^, which has been attributed to the route of administration^18^ as well as to the presence of adulterants^18,19^, leading to alterations in gene expression ^20,21^ and in the brain-gut axis ^22^, which are likely related to deficits in interoceptive processing^7^.

There is growing evidence linking cocaine consumption with a broad spectrum of neurocognitive deficits, both in occasional and chronic users ^11,23,24^. In particular, the consumption of cocaine has been consistently associated with impaired executive functions (EF), ^12,24–29^, such as working memory, cognitive flexibility, decision making and inhibitory control ^12,23,28,30–33^. Impaired attention has also been reported in chronic cocaine users, who present a slowdown in processing speed as well as difficulties in tasks that require sustained attention and set-shifting ^12,28,34^. Additionally, cognitive deficits have been associated with an early dropout of addiction treatments^35^. Specifically, cognitive flexibility and inhibitory control had been related as predictors of maintenance in treatments, as perseverative errors being a predictor for early dropout ^36,37^. Therefore, knowledge of the cognitive functioning of patients on addiction treatment has been proposed as much as significant information to predict compliance and maintenance of treatment^38^ and to adapt interventions^39^. However, several discrepancies exist, such as the absence of EF impairments suggested by a recent meta-analysis ^13^, and even improved EF reported for cocaine users ^26^. These inconsistencies could arise due to the variability in the route of administration of the drug, prompting the need for studies which specifically address neuropsychological deficits in groups of individuals addicted either to the smoked or insufflated versions of the drug.

Despite evidence suggesting that the route of administration should be taken into account to assess the short and long term effects of cocaine consumption ^1–3^, to our knowledge no study to date has characterized clinically relevant neuropsychological variables separately in populations of individuals with histories of SC and IC dependence (SCD and ICD, respectively), together with a simultaneous assessment of the structural and functional correlates of neuropsychological deficits. We focused our study on a particular form of SC, termed coca paste, an intermediate product in the extraction of cocaine hydrochloride that is especially frequent in Latin America ^16^. Coca paste presents fast pulmonary absorption and intense but short-lasting effects ^16^ and is frequently adulterated with other active compounds ^40–42^. Like other forms of SC ^10^ the consumption of *coca paste* is most widespread among adolescents in vulnerable sectors of society, as reported by the Organization of American States (OAS) and by the United Nations Office on Drugs and Crime (UNODC), and thus represents a significant public health concern.

We compared cognitive and executive function in 25 SCD, 22 ICD and 25 healthy controls (CTR), and complemented this analysis with multimodal neuroimaging data comprising functional neuroimaging recordings of spontaneous brain activity, and an assessment of grey matter density (GMd) by means of voxel-based morphometry. Finally, we combined both sets of results to investigate potential associations between changes in brain structure and function, behavioral signatures of long-term cocaine consumption, and how these associations depend upon the route of administration.

## Material and methods

### Participants

We recruited seventy-two participants, of which 25 fulfilled the DSM-IV criteria for SCD and 22 for ICD and 25 matched CTR. Participants with a history of cocaine abuse presented an early onset of drug consumption (between 14 and 16 years old) and some presented abuse and/or dependence to other drugs (see SI 1, Table 1). For a comprehensive assessment of lifetime substance use we used the Alcohol, Smoking and Substance Involvement Screening test^43^ combined with individual psychiatric interviews (see SI 1, Table S2). The SCD and ICD groups presented a significant preference for the use of SC and CC, respectively. Both groups of participants were recruited as voluntary in-patients in therapeutic communities and presented at least one year of problematic drug use. No participants presented history of neurological disease or active Axis I disorder previous to the onset of drug consumption (as evaluated with MINI-Plus, see SI 1, Table S3), nor evidence of past head injury with loss of consciousness and MRI contraindications (including pregnancy, claustrophobia, and non-removable metal in the body). Subjects under high doses of psychiatric medication were excluded (Table S4) (see SI *1, Table S4)*. Matching criteria were based on gender, age, years of education and socioeconomic status (*Table 1*). Controls were volunteers who did not have a history of drug abuse nor previous diagnosis of neurologic and/or psychiatric disorders.

**Table 1.**
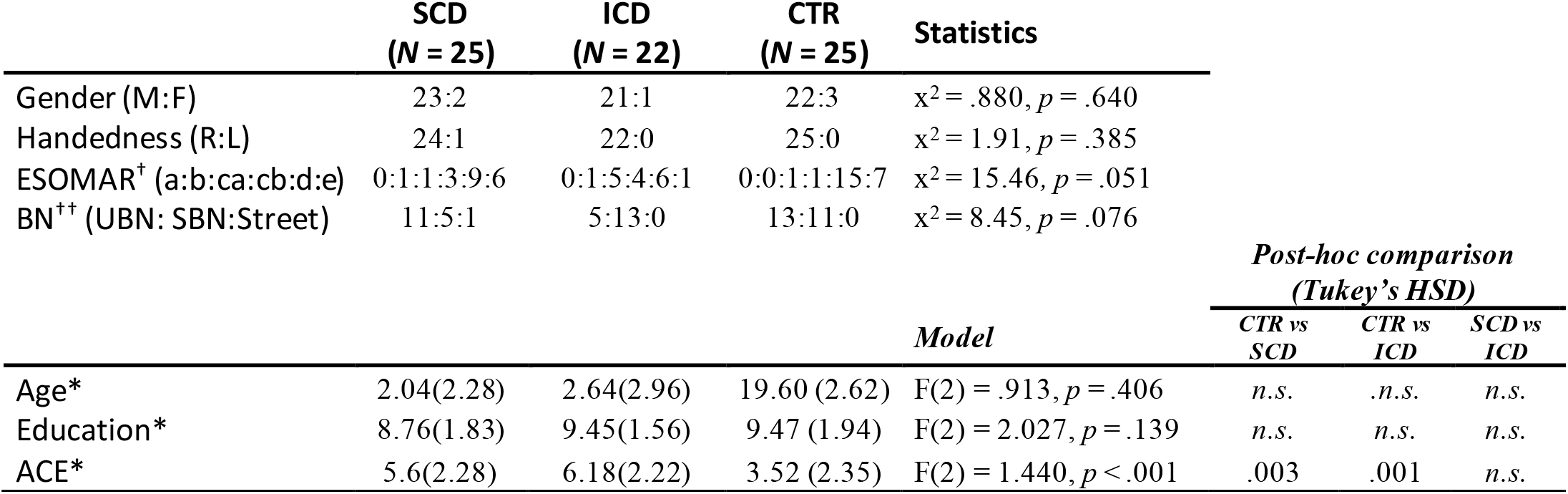
Demography: * Values are expressed as mean (standard deviation). † The European Society for Opinion and Marketing Research (ESOMAR) for socioeconomical capacity categories are very-high (a), high (b), middle-high (ca), middle (cb), middle-low (d) and low (e). †† unsatisfied basic needs (UBN), satisfied basic needs (SBN) or living in the street. CTR: Control; SCD: Smoked cocaine dependent; ICD: cocaine hydrochloride dependent.

All participants provided written informed consent in accordance with the Helsinki declaration. This study was approved by the ethics committee at Favaloro University following international principles for human medical research (N° 609/16, record 554).

### Cognitive assessment

Participants completed a thorough neuropsychological (NPS) battery assessing fluid Intelligence [Matrix Reasoning ^44^], attention [Forward Digit span, ^44^; Corsi block-tapping test ^45^; Symbol-Digit Modality ^46^; Trail Making Test Part A ^47^], executive functions [INECO Frontal Screening ^48^; Backward Digit Span ^44^; Backward Corsi block-tapping test ^45^; Letter and Number Organization ^44^; Phonological Fluency Test ^49^); Trail Making Test Part B ^47^; Hayling Test ^50^; Similarities ^44^; Modified Wisconsin Card Sorting Test ^51^], memory [Rey Auditory Verbal Learning Test ^52^; Rey-Osterrieth Complex Figure Test ^52^], language [Vocabulary ^44^; Semantic Fluency ^49^] and social cognition [Reading the Mind in the Eyes Test ^53^; Reading the Mind in the Voice ^54^; MINI-SEA ^47^; The Awareness of Social Inference Test ^55^]. A detailed explanation of each task and their scoring can be found in SI 2.

### MRI image acquisition and preprocessing

We analyzed 65 MRI recordings (20 SCD, 22 ICD and 23 CTR), omitting participants who either abandoned the protocol or opted out of the MRI session (for details see SI 3). Participants were scanned in a 1.5 T Phillips Intera scanner with head coil SENSE-NV, phased array with 8 channels for structural T1 scans, and functional GRE-EPI images (details on SI 3, Table S7). During resting-state the participants were instructed to avoid thinking about anything in particular and to keep their eyes closed without falling asleep during 10 minutes ^56–58^. For functional image analysis we followed the procedure outlined in previous reports from our group ^7,59^ in which images were preprocessed using the Data Processing Assistant for Resting-State fMRI (DPARSF V4.3) ^60^. Structural images were preprocessed with the CAT12 Toolbox from Statistical Parametric Mapping 12 software package (https://www.fil.ion.ucl.ac.uk/spm/) ^61^ and analyses were carried out using total intracranial volume (TIV) as a variable of no interest following previous reports^7^ (detailed explanation in SI 3).

## Statistical Analysis

### Cognitive Assessment

To compare behavioral performance between groups we used non-parametric Kruskal-Wallis tests with Wilcoxon-Mann-Whitney tests for post hoc comparisons, corrected by False Discovery Rate (FDR) with a significance level given by *p*-FDR corrected <.05. The choice of non-parametric statistics was motivated by the heterogeneous distributions seen in different neuropsychological variables.

The performance of each individual task involves multiple neurophysiological processes^14^. To extract meaningful information from the multivariate dataset of neuropsychological evaluations we used a use an exploratory factorial model based on Varimax rotation ^62^. Previous to this analysis we inverted the scores of variables for which better performance implied worse scores and excluded neuropsychological domains showing significant effects in only one task (see SI 2). To establish the number of relevant factors we explored latent domains using a graph representation where individual nodes represent NPS tasks and edges represent the Spearman correlation value between the corresponding tasks. We then determined the optimal modular structure using the Louvain agglomerative algorithm^63^ giving 3 modules (see results and SI 2). The individual scores of the resulting factors were calculated as the loading projection of the original variables. To confirm the factor structure, we compared group scores with non-parametric Kruskal Wallis tests with Wilcoxon-Mann-Whitney tests for post hoc comparisons, reported using a significance level of *p*-FDR corrected <.05.

To explore the relationship between performance and information related to drugs use, we fitted a random forest regression model for each of the factor scores using consumption data as the dependent variable (see SI 2, Table S5 and SI 3, Table S6). We reported the mean/standard deviation (SD) of R^2^ and the associated *p*-values. We repeated this analysis using 1000 iterations with randomly permutated target values, and we obtained the associated p-value as the proportion of R^2^ values with permutation that exceeded the mean R^2^ without permutation. Results were considered significant at a threshold of *p* < .05.

### Brain-behavior correlation analysis

Beside alterations specific to the striatum and related structures^64,65^, there is ample variability in previous reports of GMd changes in chronic users of cocaine^66^. Also, our sample comprised young adults with a history of abuse but not currently engaged in cocaine consumption. Given these uncertainties we opted for an exploratory whole-brain analysis, expecting GMd changes located in the striatum but potentially also in other brain regions. We first compared GMd between consumer groups and CTR using t-tests (whole-brain analysis, *p* < .001 uncorrected, extent threshold = 50 voxels [García-Cordero et al., 2016] to confirm differences at the striatum and related structures^64,65^. TIV was used as a covariate to discard the influence of brain-size differences. We extracted GMd values from the ROIs on the caudate for each whole-brain comparison and subject. We assessed the association between GMd and Factor 1 scores via Spearman correlation, with statistical significance set at at *p*-FDR corrected < .05, and excluding subjects as outliers whenever their GMd or factor score were 2 SDs above the group’s mean.

To examine associations between functional connectivity (FC) and behavior, we extracted the Blood Oxygen Level Dependent (BOLD) signals from the two left caudate ROIs corresponding to the significant clusters found in the VBM analysis, and correlated them with the BOLD signals from every voxel in the brain using Pearson’s correlation, resulting in a whole-brain map of Fisher-transformed z-values corresponding to the FC with each seed ROI. We used SPM’s multiple regression module to explore associations between behavioral scores and FC maps of SCD (whole-brain analysis, *p* < .05-cluster family wise corrected, extent threshold = 50 voxels). To test differences in connectivity between clusters associated with significant changes in performance, we extracted FC values and compared them between groups using non-parametric Kruskal-Wallis tests with Wilcoxon-Mann-Whitney tests for post hoc comparisons, considered significant at *p*-FDR corrected <.05.

Finally, to evaluate whether differences in GMd (obtained using TIV as covariate) could account to the first factor of the behavioral differences, mediation analysis was performed. We selected ROIs from the significant clusters determined by the FC multiple regression analysis, and performed a mediation analysis with behavioral (Factor 1), structural (GMd of the left caudate ROI) and functional (FC of the left caudate ROI) variables. Outliers were determined as samples 2 SD above the mean. All assumptions of regression were confirmed. To perform the mediation analysis we followed the steps described by Baron and Kenny ^67^, applying a bootstrap procedure with 5000 repetitions (p <.05) to assess the contribution of FC and GMd to Factor 1 scores.

## Results

### Cognitive Assessment

Subjects with history of cocaine abuse showed poorer performance in a fluid Intelligence task (Matrix Reasoning subtest) presenting lower scores than CTR (SCD, *p* <.001; ICD, *p* <.001; Table 2, Figure 1A). Differences in a*ttention* were found between groups, where the SCD group exhibited a lower score than ICD and CTR in the Digits forward span task (ICD, *p* = .011; CTR, *p* = .010), Symbol-Digit Modality (ICD, *p* = .0.21; CTR, *p* = .002) and TMT-A (ICD, *p* = .006; CTR, *p =* .013). In the Corsi block-tapping test, both the SCD and the ICD group exhibited a lower score than CTR (*p* < .001 and *p* = .05 respectively). (Table 2, Figure 1).

**Table 2.** Cognitive performance: Values are expressed as mean (standard deviation. N). SCD = Smoked cocaine dependent; ICD = cocaine hydrochloride dependent; CTR = Control; FI = Fluid Intelligence; A = Attention; EF = Executive Functions; M = Memory; L= Language; SC Social Cognition; TMT A= Trail Making A; TMT B= Trail Making B; WCST= Wisconsin Card Sorting Test; LNST= Letter-Number Sequencing Test; IFS: INECO Frontal Screening; RAVLT: Rey Auditory Verbal Learning Test; RCF= Rey Complex Figure; RMET=Reading the Mind in the Eyes test; RMVT=Reading the Mind in the Voice test; TASIT= The Awareness of Social Inference Test

|  |  |  |  |  |  | Mann-Whitney p-FDR corrected |  |  |
| --- | --- | --- | --- | --- | --- | --- | --- | --- |
|  |  |  |  |  |  | SCD vs CTR | ICD vs CTR | SCD vs ICD |
|  |  | SCD (25) | ICD (22) | CTR (25) | Model |  |  |  |
| **FI** | Matrix Reasoning (WAIS III) | 6.24 (2.59, 25) | 6.50 (2.62, 22) | 9.60 (2.57, 25) | KW(2,72)=19.066, p < .001 | < .001 | < .001 | n.s |
| **A** | Digits forward | 4.72 (1.02, 25) | 5.45 (.80, 22) | 5.52 (1.12, 25) | KW(2,72)=8.796, p = .012 | .010 | n.s | .011 |
|  | Corsi block-tapping test | 5.21 (.66, 24) | 5.55 (.96, 22) | 6.16 (1.18, 25) | KW(2,71)=1.751, p = .005 | <.001 | .050 | n.s |
|  | TMT A (seconds) | 39.64 (1.50, 25) | 31.41 (9.60, 22) | 33.08 (11.92, 25) | KW(2,72)=9.416, p = .009 | .013 | n.s | .006 |
|  | Symbol-Digit | 4.32 (12.45, 25) | 41.91 (7.90, 22) | 48.56 (8.31, 25) | KW(2,72)=1.495, p = .005 | .002 | .021 | n.s |
| **EF** | Digits backwards | 3.48 (.77, 25) | 3.77 (.69, 22) | 3.92 (.95, 25) | KW(2,72)=8.796, p = .012 | n.s | n.s | n.s |
|  | Backwards Corsi block tapping | 4.54 (1.14, 24) | 4.45 (1.01, 22) | 5.08 (1.41, 25) | KW(2,71)=2.969, p = .227 | n.s | n.s | n.s |
|  | TMT B (seconds) | 11.20 (5.08, 25) | 103.23 (72.92, 22) | 77.08 (26.29, 25) | KW(2,72)=9.098, p = .011 | .003 | n.s | n.s |
|  | Phonological Fluency | 1.36 (3.34, 25) | 14.73 (4.03, 22) | 13.04 (4.68, 25) | KW(2,72)=12.224, p = .002 | .033 | n.s | <.001 |
|  | Hayling Test | 12.12 (8.60, 25) | 9.55 (7.73, 22) | 4.68 (6.03, 25) | KW(2,72)=12.033, p = .002 | .001 | .026 | n.s |
|  | WCST - categories | 4.00 (1.96, 25) | 3.91 (1.57, 22) | 5.04 (1.31, 25) | KW(2,72)=6.076, p = .048 | .049 | .024 | n.s |
|  | WCST - errors | 7.08 (5.46, 25) | 9.64 (4.10, 22) | 5.36 (4.25, 25) | KW(2,72)=10.610, p = .005 | n.s | <.001 | .041 |
|  | WCST - perseverations | 3.49 (3.49, 25) | 3.95 (2.66, 22) | 1.76 (2.33, 25) | KW(2,72)=9.353, p = .009 | n.s | .002 | n.s |
|  | LNST (WAIS III) | 4.80 (2.74, 25) | 6.45 (2.72, 22) | 7.88 (2.86, 25) | KW(2,72)=13.615, p = .001 | < .001 | n.s | n.s |
|  | Similarities (WAIS III) | 5.76 (2.07, 25) | 6.55 (1.65, 22) | 7.52 (2.10, 25) | KW(2,72)=11.193, p = .004 | <.001 | n.s | n.s |
|  | IFS | 18.54 (3.23, 13) | 19.67 (5.78, 15) | 23.48 (2.75, 25) | KW(2,53)=18.036, p < .001 | < .001 | .008 | n.s |
| **M** | Immediate (RAVLT) | 43.44 (8.97, 25) | 46.82 (7.50, 22) | 51.04 (6.79, 25) | KW(2,72)=1.677, p = .005 | <.001 | n.s | n.s |
|  | Delayed (RAVLT) | 1.36 (2.78, 25) | 9.55 (3.25, 22) | 12.28 (1.74, 25) | KW(2,72)=11.215, p = .004 | .013 | .002 | n.s |
|  | Recognition (RAVLT) | 12.60 (1.85, 25) | 13.32 (1.36, 22) | 13.68 (1.38, 25) | KW(2,72)=5.077, p = .079 | n.s | n.s | n.s |
|  | Immediate (CFR) | 3.68 (2.81, 25) | 31.84 (1.66, 22) | 32.08 (2.84, 25) | KW(2,72)=4.244, p = .120 | n.s | n.s | n.s |
|  | Delayed (CFR) | 19.14 (6.29, 25) | 2.00 (4.46, 22) | 22.38 (5.83, 25) | KW(2,72)=3.744, p = .154 | n.s | n.s | n.s |
| **L** | Semantic Fluency | 17.68 (5.25, 25) | 2.00 (4.46, 22) | 18.84 (3.36, 25) | KW(2,72)=3.046, p = .218 | n.s | n.s | n.s |
|  | Vocabulary (WAIS III) | 5.96 (1.79, 25) | 7.27 (1.35, 22) | 7.40 (2.24, 25) | KW(2,72)=9.457, p = .009 | .004 | n.s | .021 |
| **SC** | Mini SEA | 22.00 (4.29, 25) | 22.10 (4.26, 22) | 22.64 (3.54, 25) | KW(2,72)=.680, p = .712 | n.s | n.s | n.s |
|  | RMET | 1.00 (2.25, 25) | 1.00 (2.25, 22) | 11.12 (1.83, 25) | KW(2,72)=4.564, p = .102 | n.s | n.s | n.s |
|  | RMVT | 12.40 (2.55, 25) | 13.86 (1.67, 22) | 14.04 (2.42, 24) | KW(2,71)=7.368, p = .025 | .018 | n.s | .021 |
|  | TASIT | 8.17 (1.55, 24) | 8.73 (.94, 22) | 8.71 (.95, 24) | KW(2,70)=1.917, p = .383 | n.s | n.s | n.s |

**Figure 1.**
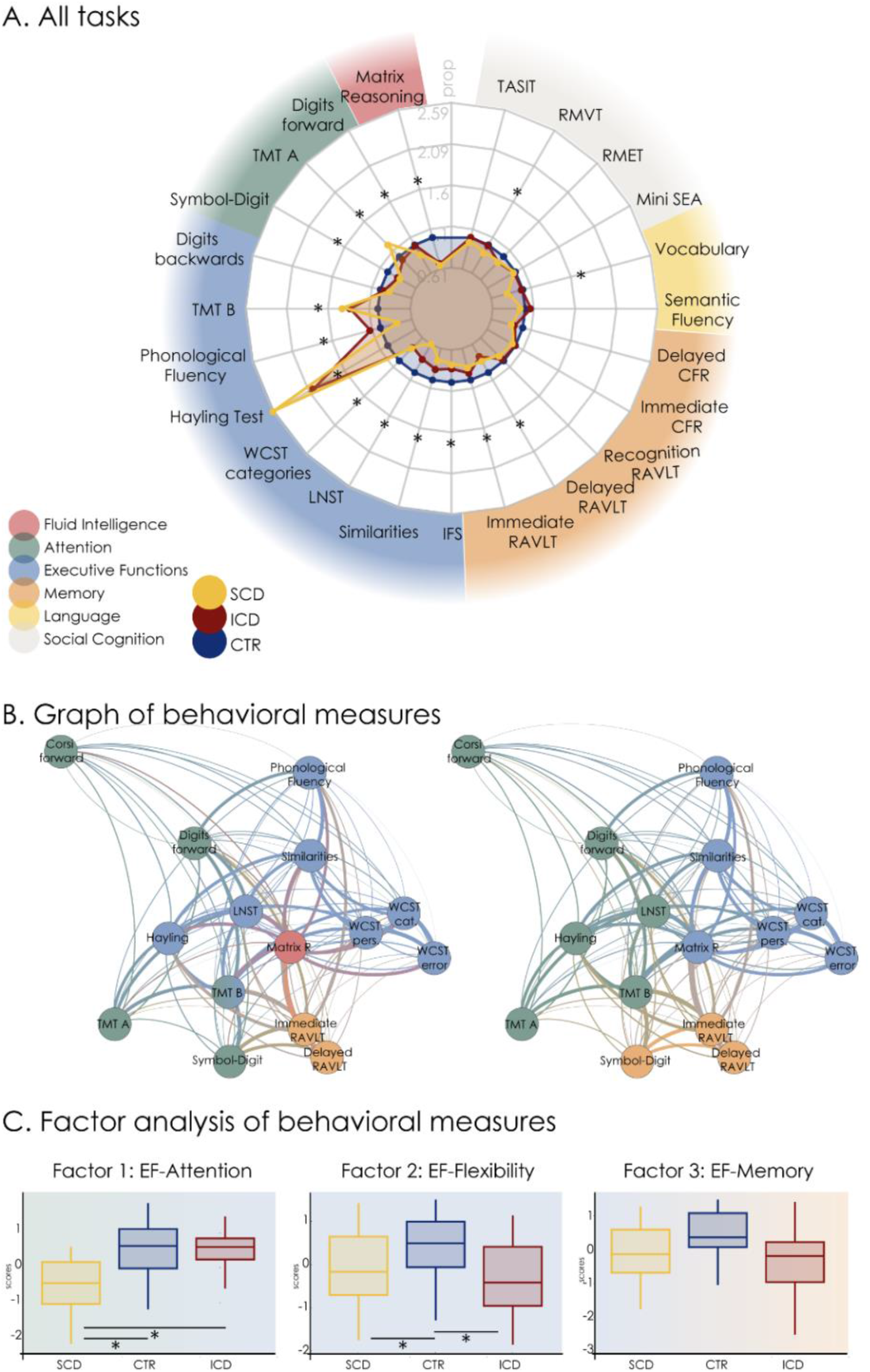
Behavioral performance: A. Comparison of the performance of each group across neuropsychological tasks by domain. The performance of controls (blue circle) is taken as reference. B. Graphs of task performance similarity, colored by domain (Left) and by community structure (Right). In each of these graphs the nodes represent a single task and the edges connecting them represent the correlation between the associated performances (i.e. the closer the nodes, the higher the correlation). C. Factor analysis of neuropsychological measures. The 3 first factors are named by their load on individual tasks. Yellow indicates the Smoked Cocaine Dependent (SCD)group; blue indicates healthy controls (CTR); and red indicates the IC dependent (ICD) group. *indicates significant differences (p-FDR corrected <0.05). EF= Executive Functions; TMT A= Trail Making A; TMT B= Trail Making B; WCST= Wisconsin Card Sorting Test; LNST= Letter-Number Sequencing Test; IFS: INECO Frontal Screening; RAVLT: Rey Auditory Verbal Learning Test; RCF= Rey Complex Figure; RMET=Reading the Mind in the Eyes test; RMVT=Reading the Mind in the Voice test; TASIT= The Awareness of Social Inference Test

*In the executive function* domain, significant differences between the groups in the performance of several tasks were found. There were significant differences with lower performance for consumers groups relative to CTR on the Hayling Test (SCD, *p* < .001; ICD, *p* = .026), in the total categories score of the WCST (SCD, *p* = .049; ICD, *p* = .024), in the IFS total score (SCD, *p* < .001; ICD, *p* = .008). However, in WCST the ICD group showed a significantly higher number of perseverations (*p* = .002) and errors (*p* = .001) throughout the task in comparison with CTR. The SCD group exhibited a lower performance than CTR in the TMT-B than CTR (*p* = .003), Similarities (*p* < .001) and Number Sequencing subtest (*p* < .001). Furthermore, differences were also found in the Phonological Fluency test, with SCD exhibiting a lower performance than ICD (*p* < .001) and CTR (*p* = .033). No significant differences were found between the groups in the performance of the Digits backwards span task and the Backwards Corsi block-tapping task (Table 2, Figure 1).

We also found differences in *memory* tasks where SCD exhibited a lower performance in the immediate recall tasks (RAVLT) than CTR (*p* < .001). Moreover, on the delayed phase (RAVLT) both SCD (*p* = .013) and ICD (*p* = .002) showed a lower performance than CTR. No significant differences were found between groups in the recognition phase (RAVLT) nor for the acquisition phase and the delayed recall phase of the visual memory task (CFR). In *language* tasks, the SCD group exhibited a lower performance on the Vocabulary subtest compared both to ICD (*p* = .021) and control groups (*p* = .004). Regarding *social cognition*, only the SCD group exhibited a lower performance on Reading the Mind in the Voice than both ICD (*p* = .021) and CTR (*p* = .018). No significant differences were found between the groups in any other task (Table 2, Figure 1).

Figure 1B shows the graphs obtained after correlating the tasks for which we found significant differences between SCD/ICD and CTR by their performance vectors. Panel B (right) is coloured according to the modular assignment, and panel B (left) is coloured according to the neuropsychological domains. The optimal modularity of this graph was 0.11, with 3 modules representing 40%, 40% and 20% of all tasks, respectively. The graph coloured according to the domains illustrates the separation between executive functions, attention and memory.

Table 3 shows an exploratory factor analysis with three factors (EF-Attention, EF-Flexibility and EF-Memory). In accordance with the modularity structure, the first two factors comprised mainly attention and flexibility tasks, and the last comprised memory tasks; also, matrix reasoning showed a similar association with each factor. When comparing groups among factors we found (Figure 1, panel C) differences between the group scores of Factor 1 (FE-Attention), where the SCD group exhibited a lower score than ICD (p < .001) and CTR (p < .001); Factor 2 (FE-Flexibility), where both the SCD group (p = .041) and the ICD group (p = .009) exhibited a lower score than CTR. On Factor 3 (FE-Memory), the ICD group exhibited a non-significant trend towards lower scores than CTR (See Table 4 and Figure 1). As Factor 1 distinguished among groups, we centered our analysis in this EF-Attentional factor to further explore brain-behaviour relationship.

**Table 3.**
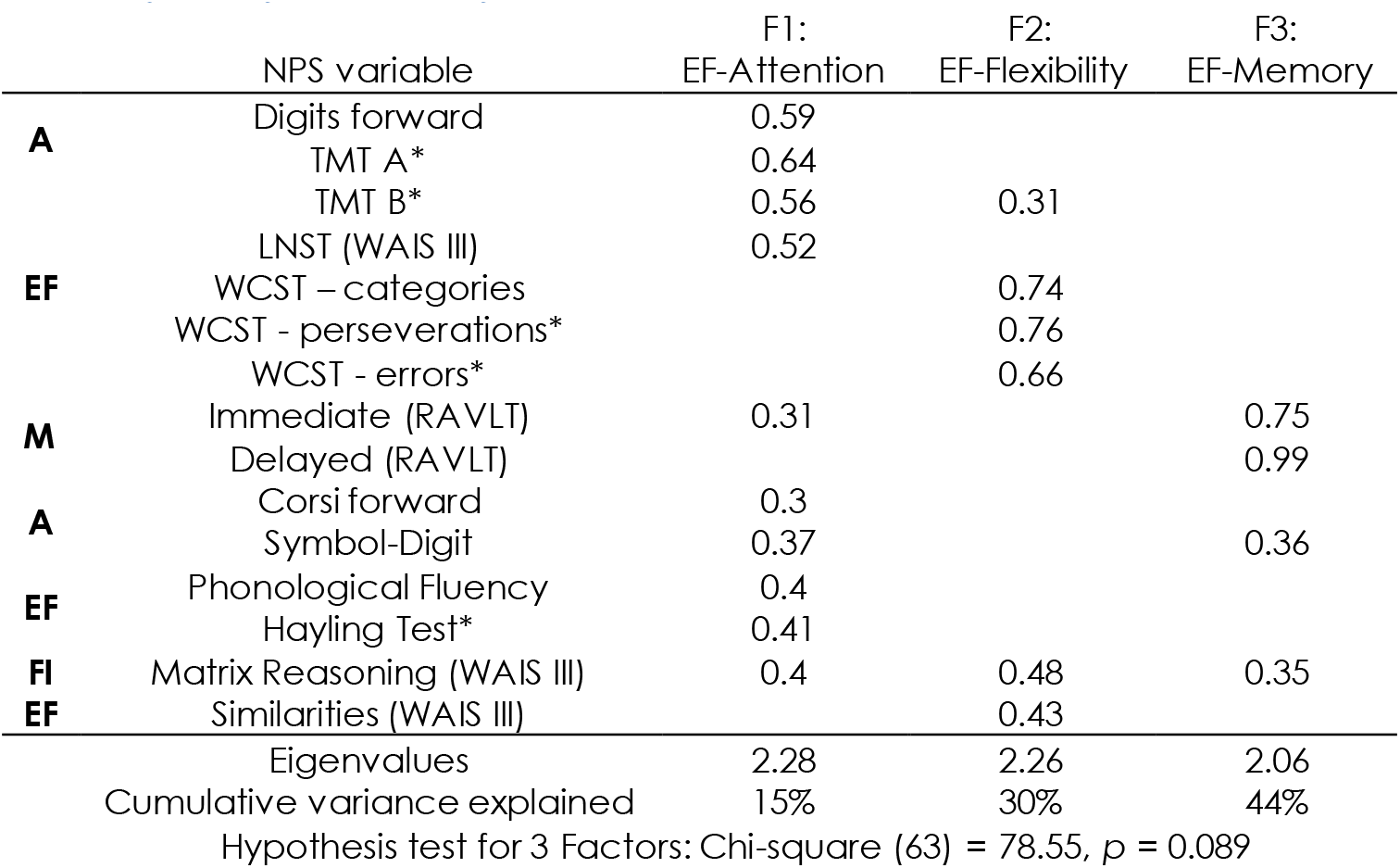
Exploratory factorial analysis: *Variables inverted before factor analysis; FI = Fluid Intelligence; A = Attention; EF = Executive Functions; M = Memory; L= Language; SC Social Cognition; TMT A= Trail Making A; TMT B= Trail Making B; WCST= Wisconsin Card Sorting Test; LNST= Letter-Number Sequencing Test

**Table 4.**
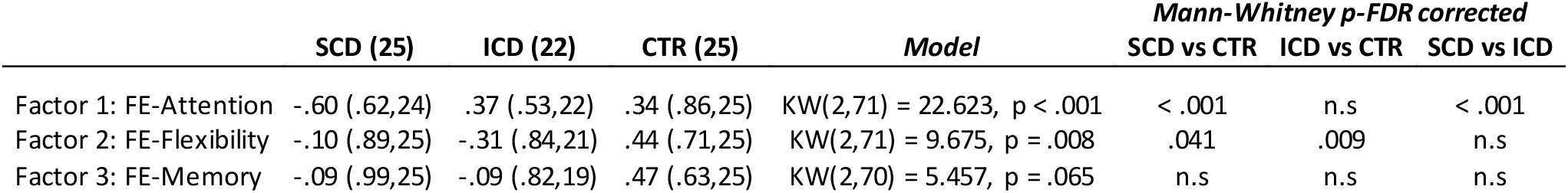
Factor statistics: Values are expressed as mean (standard deviation, N). SCD = Smoked cocaine dependent; ICD = cocaine hydrochloride dependent; CTR = Control.

Using Random Forest regression we found a significant association between drug polyconsumption data and Factor 1-Attention (R^2^ = 0.201/0.013, *p* = 0.001/0.002, mean/sd values), significantly different from permuted data (*p* < 0.05, 1000 permutations), mostly due age and THC age onset (see SI 2, Figure 1 and SI Table 5).

### Brain-Behavior relationship analysis

The SCD group presented reduced GMd in right middle frontal and left caudate regions relative to CTR. The ICD group showed reduced GMd in occipito-parietal, right middle frontal and superior motor areas relative to CTR. ICD also presented hypertrophic parahippocampal-fusiform regions bilaterally compared to the CTR group. Comparisons between groups of cocaine users showed that SCD presented reduced GMd over bilateral caudate relative to ICD, but the opposite was observed at occipito-parietal and medial frontal regions (see Figure 2 and Table S8). In summary, the only region that distinguished SCD from CTR and ICD was the left caudate.

**Figure 2.**
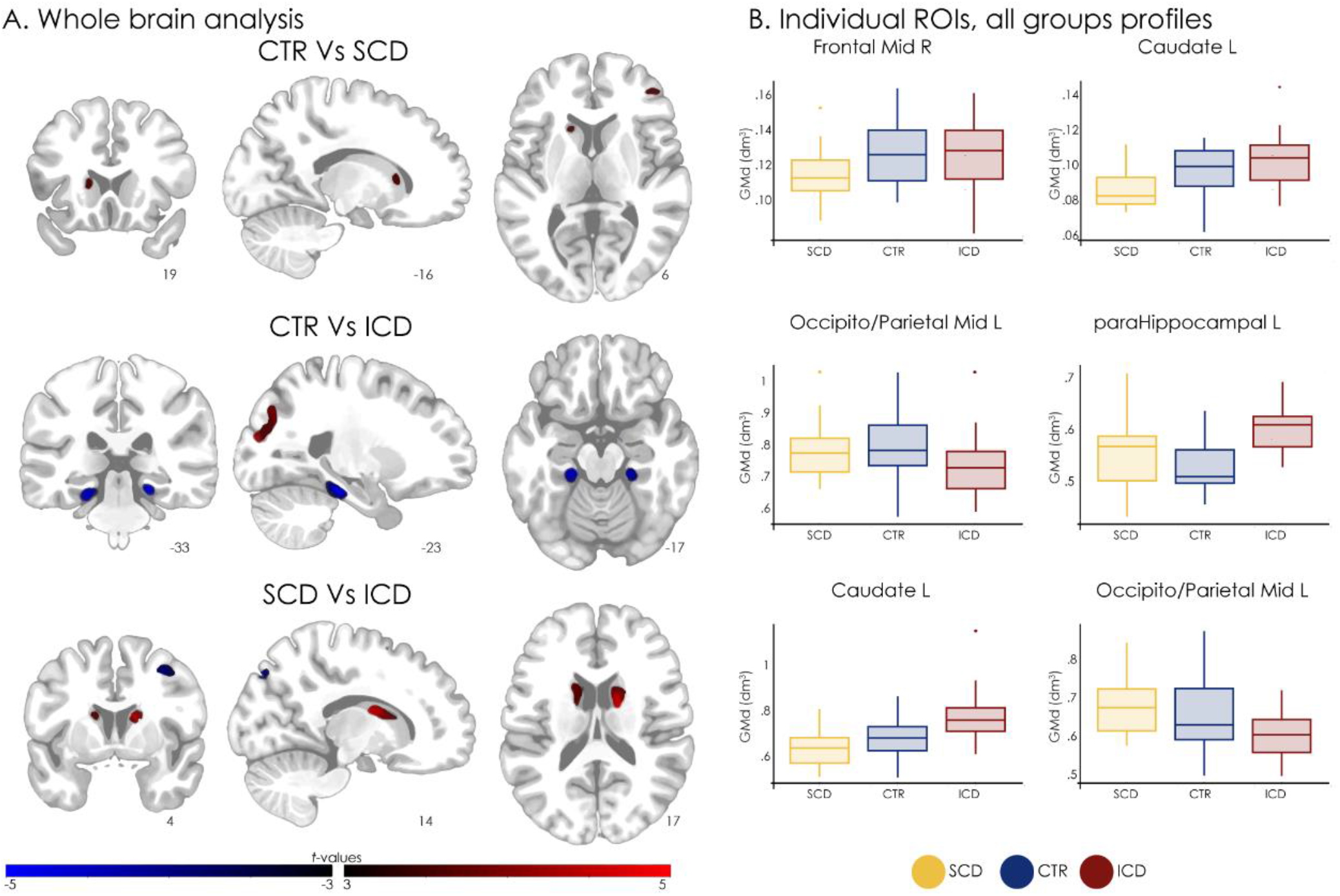
GMd results: A. Explorative comparison of the GMd between groups. SCD showed reduced GMd in right middle frontal and left subcortical caudate regions, and ICD showed reduced GMd in occipito-parietal, right middle frontal and superior motor areas relative to CTR. ICD also presented hypertrophic parahippocampal-fusiform regions bilaterally. CTRs showed reduced GMd compared to ICD in medial temporal areas. SCD showed reduced GMd in bilateral caudate, and higher GMd in occipito-parietal and medial frontal regions compared to ICD (p < .001 uncorrected, extent threshold = 50 voxels). B. GMd extracted from the two largest significant regions in each whole-brain comparison. Yellow indicates the Smoked Cocaine Dependent (SCD) group; blue indicateshealthy controls (CTR); and red indicates the IC dependent (ICD) group.

Left caudate regions presented a significant association with Factor 1 (EF-Attention) only for SCD but not for CTR nor ICD, with less gray matter implying better performance (see Figure 3 and Table 5). FC of caudate regions did not differ among groups (Table 6). Moreover, higher FC between bilateral frontal cortex and left caudate predicted worst performance only for SCD, while CTR did not show such an association (FC between inferior frontal ROIs and the dorsal caudate was nearly 0, and independent of Factor 1). For high performances, the inferior frontal-dorsal caudate FC of CTR approached that of SCD.

**Figure 3.**
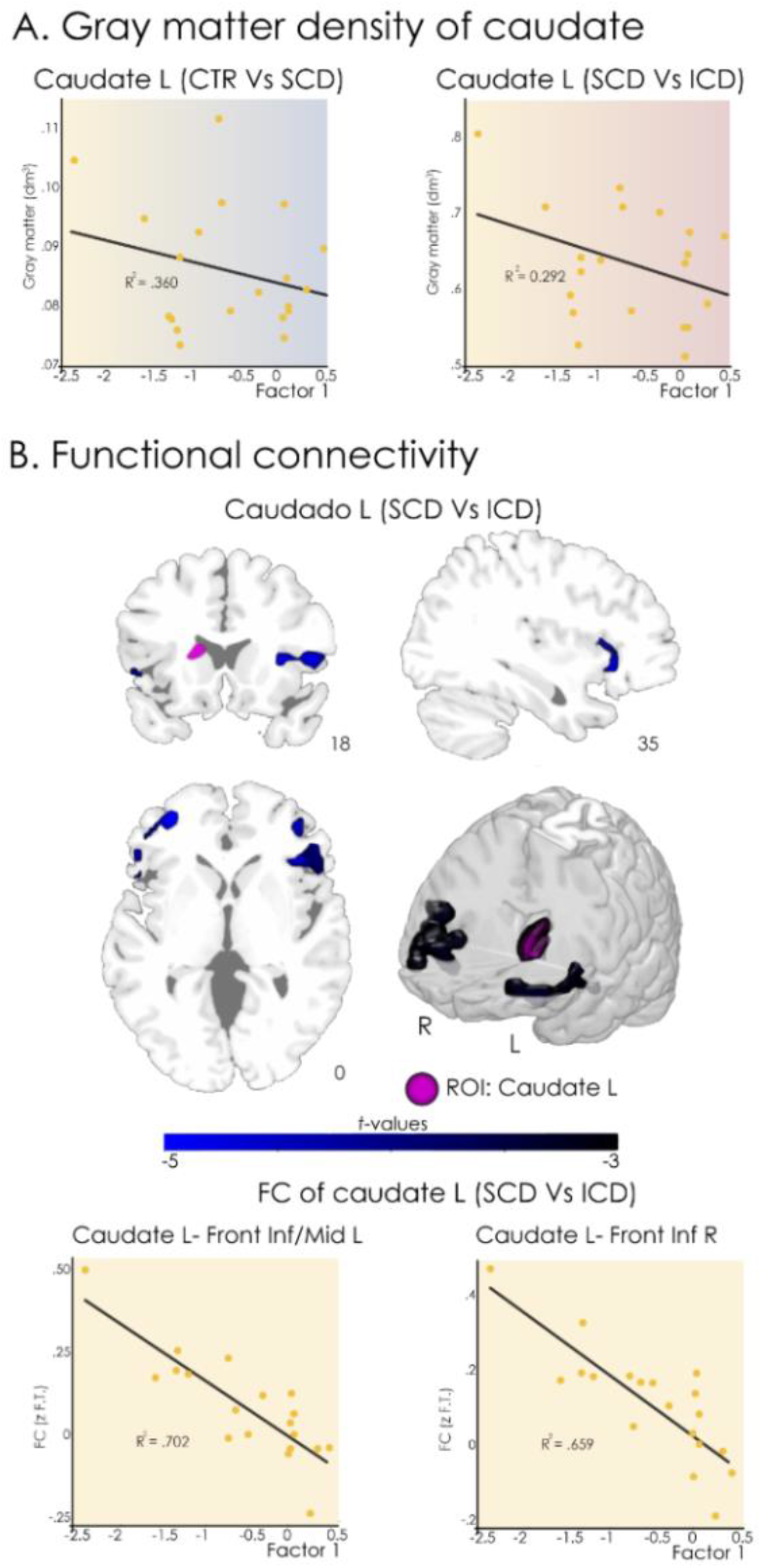
Factor 1: Structural and functional associations in SCD: A. Comparison of grey matter density of caudate in association with factor 1 (EF-Attention) between groups, only in SCD left caudate regions presented a significant association with Factor 1: (p < .001 uncorrected, extent threshold = 50 voxels). B. Functional connectivity between SCD and factor 1 (EF-Attention), higher FC between bilateral frontal cortex and left caudate predicted worst performance.

**Table 5.**
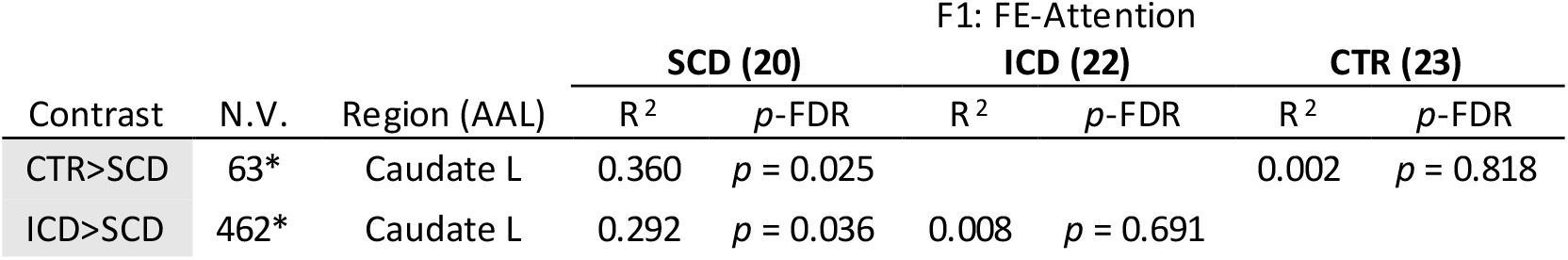
Factor 1 association with caudate ROIs GM: SCD = Smoked cocaine dependent; ICD = cocaine hydrochloride dependent; CTR = Control.

**Table 6 FC.**
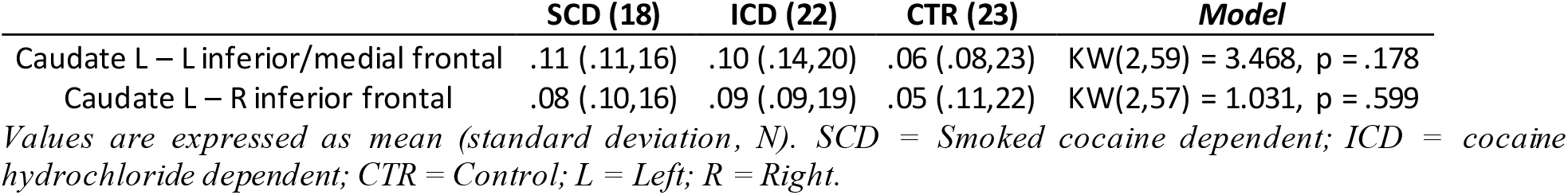
Values are expressed as mean (standarddeviation, N). SCD = Smoked cocaine dependent; ICD = cocaine hydrochloride dependent; CTR = Control; L = Left; R = Right.

Only for SCD both structural and FC of left caudate ROI was associated with task performance. We then analyzed whether the relationship was driven by FC. Mediation analysis revealed that partialling out the variance of FC between left caudate and left frontal-inferior or right frontal medial ROI significantly attenuated the relationship between GMd and Factor 1 scores (“c’ pathway”, Left: t(15) = −2.10, p = 0.255; Right: t(15) = −.34, p = 0.863, see SI 4, Figure S2). Statistics for left: Bootstrapping average indirect pathway, b = −3.725 [95% CI, -7.931 to -0.331], Sobel test: Z = −1.97, p = .048, statistics for right: Bootstrapping average indirect pathway, b = −5.487 [95% CI, -10.003 to -1.646], Sobel test: Z = −2.60, p = .009.

## Discussion

Despite vast previous literature on the behavioral and neurophysiological consequences of long-term cocaine addiction, both insufflated and smoked cocaine are frequently investigated as parts of a single category, and as a result little is known about the interaction between neurophysiological and neuropsychological variables and the route of administration of the drug ^10,12^. In the present study we used clinically relevant neuropsychological tasks spanning multiple domains in combination with structural and functional imaging to shed light on potential differences in the deficits associated with smoked and insufflated cocaine. We found that different routes of administration led to equally different profiles of neurocognitive impairment, with the SCD group being specifically associated with deficits in attention and executive functions. We also found that those profiles were consistent with the neurophysiological data, with close associations between structural, functional and behavioural variables for the SCD group. Taken together, our results support the clinical observation that smoked forms of cocaine are associated with deeper neuropsychological impairments and higher abuse potential ^4–7^.

As consistently shown by previous studies ^12,24,27,28^, we found that the SCD and ICD groups exhibited EF impairments in comparison with CTR. Here, we specifically addressed the route of administration, showing that subjects in the SCD group exhibited overall more profound deficits compared to the ICD group. Regarding *memory*, we found that performance was more pronouncedly impaired in SCD than in ICD, but deficits were seen in both groups specifically in the executive components of memory (immediate and delayed RAVLT). However, we did not find significant differences in the recognition phase of the memory task, as opposed to previous studies ^68,69^ that failed to account for potential differences related to the route of administration. Finally, in our matched sample we only found differences between groups in one social cognition task (Reading the mind in the voice), which is consistent with previous literature showing that the aggressive behaviour observed in individuals with history of smoked cocaine abuse could be mostly driven by the socioeconomic factors surrounding drug use ^70^.

All the tasks we assessed in the present study imply multiple domains, here explored by means of graph and factor analyses. It is important to highlight that all variables showing differences between groups were the most representative to measure executive functions, independently of the original domains. When comparing the distribution of modules based on performance correlations (Figure 1, left panel), we found that the modules mainly overlapped with the classical domains, with the following exceptions: Symbol-Digit task (a task with a significant memory component ^71^) fell in the “memory” cluster and TMT B, LNST and Hayling fell into “attention” cluster. Matrix reasoning fell in the “executive” cluster, but was closely related to all other tasks. Overall, factor analysis showed that both groups presented similar impairments in EF-Flexibility. However, while the ICD group presented a trend towards the impairment of memory-related processing, then SCD group showed a stronger attentional deficit. The relative importance of *executive, attentional* and *memory* domains has been controversial in the neuropsychological study of cocaine consumption ^12,24,27,28^. We suggest that these inconsistencies could arise due to incomplete matching ^10^ of critical variables (such as SES), polyconsumption^13^, and most critically by differences in the route of administration^12,24,27,28^, possibly the most under-investigated factor in the current literature.

While we would expect that if the cognitive failures were driven by cocaine consumption data, frequency or amount of SC od IC should predict performance. Instead, our results show age and THC age onset as the mainly relevant variable to explain attention. The populations we investigated presented an early onset of drug abuse, and cocaine consumption during adolescence (i.e. starting before 18 years) contributing with more profound cognitive impact relative to cocaine abuse beginning during adulthood ^27,29^. Our result suggests that EF-Attention deficits could represent a risk factor towards developing addiction to drugs, particularly during the adolescence, consistent with longitudinal studies of drug consumption initiation, suggesting that poor development and certain individual cognitive traits can predict early susceptibility to addiction (i.e. “risk model”) ^72^.

At the structural level, the pattern of behavioral deficits was consistent with group differences in GMd located at key *EF, memory* and *attention* areas. These areas have been previously implicated in the long-term effects of stimulant abuse, which induce structural alterations in regions such as the left middle frontal gyrus, hippocampus, occipito-parietal cortex, as well as the caudate nucleus ^65^. As shown in Figure 2, the GMd analysis established that frontal and caudate ROIs could distinguish SCD, while occipito-parietal and hippocampal ROIs selectively distinguished the ICD group and CTRs. Particularly, the left caudate ROI distinguished between both groups of consumers, presenting reduced GMd for SCD and hypertrophic features for ICD. Even there is an agreement over striatum physiological roll, suggesting early involvement the primary target of the drug (accumbens) evolving to habit-forming and goal-directed striatum zones such as the dorsal striatum^73^ time after the cease of consumption, previous studies of chronic cocaine consumers that were non-specific with regards to the route of administration have reported alterations in the caudate, both reduced^65^, hypertrophic^74^ or regional dependent^64^. Given that the caudate emerged as the main region that distinguished between SCD and ICD which share primary target of cocaine, and given that both groups were matched by their sociodemographic variables and history of substance abuse, we believe that the differences between groups could be attributed to the route of administration.

However, several not mutually exclusive reasons could drive the group differences found in the dorsal striatum (S.I.5, S Discussion), which are relevant in the discussion of attentional deficits. Our results show that age was the main variable in the prediction EF-Attention deficits. The striatum still undergoes development during adolescence ^75–78^, leading to improvements in cognitive performance up to levels that are characteristic of early adulthood ^77^. The age of onset of cocaine use is a strong predictor of abuse ^79^ and of the development of cognitive disorders^25^. This period of life may constitute a window of vulnerability due to the interactions with developmental factors (“risk model”) as well as due to the specific effects of dopaminergic stimulants (“exposure model”) ^72^. Our results give support to the risk model, since deficits in the EF-Attention factor could not be explained by variables associated with cocaine consumption, but were instead predicted by age and onset of THC consumption, consistently with longitudinal studies of cannabis abuse ^72,80^. Further longitudinal studies specific to individuals with problematic use of dopaminergic stimulants are needed to corroborate these findings.

The pattern of FC changes was informative of the processes underlying our neuropsychological results. Only for SC we found that the decoupling between bilateral frontal regions and the caudate explained changes in behavioral outcomes, outperforming the correlation observed with structural alterations. The association between frontal-caudate FC and EF-Attention is consistent with a neuroadaptive process similar to that already reported for insular FC and interoceptive performance^7^. Studies using animal models have shown decreased caudate-frontal functional connectivity under the acute effects of intravenous cocaine self-administration ^81,82^. Also, frontal-striatal FC predicts cocaine intake when access to cocaine is provided ^83^. This could represent a pre-existing signature that predisposes the individuals towards cocaine abuse ^84,85^.

An important limitation of our study is the relatively small sample size, which is nevertheless comparable to that of other previous studies ^85,86^. We addressed this limitation by including a control sample matched in terms of sociodemographic variables and polysubstance use, which represents a common potentially confounding factor in groups of cocaine users ^87^. We obtained groups that only differed in the preferred drug route of administration (SC or IC), a relevant factor that has been frequently neglected in the previous literature ^12,14,31^. Although our groups were matched by gender, most of the enrolled subjects were male. This limitation is shared with most studies of stimulant addiction ^88^, partially given that the majority of stimulant users are male (UNODC reports ^89^). This difficulty is heightened in the case of SC users ^40^ given that treatment options are harder to access and sustain for women ^40^. These limitations reflect the fact that our sample was obtained from a real-world clinical population recruited from multiple communities of drug users, which at the same time adds to the general validity of our results. Finally, future studies should explore whether the neurophysiological deficits are present previous to the onset of consumption, and thus whether they can predict vulnerability towards developing problematic use of stimulants.

Our results strongly support the relevance of the route of administration in the assessment of neurophysiological and behavioral impairments as a consequence of long-term cocaine abuse. We showed that different routes resulted in distinct neuropsychological profiles, and even while both groups presented EF deficits, SC consumption was associated with deeper neuropsychological impairments particularly over EF-Attention domains. We showed the convergence of these results with neurophysiological variables, establishing that the SCD group presented reduced GMd in key EF and attention areas. These findings could have important clinical implications for the understanding of differential behavioral patterns of SCD and IC and the consequent development of successful treatment strategies. Knowledge of the cognitive profile provides valuable information concerning cognitive strengths and weaknesses that are important in a clinical context ^36,38,90^. In this way, improving our knowledge of the neuropsychological deficits associated with specific routes of administration could help identify better pharmacological and psychotherapeutic interventions ^90,91^. Future longitudinal studies should investigate whether the EF-Attention deficits observed in individuals with history of problematic SC consumption constitute a marker of vulnerability (risk model), a result of the early exposure to a dopaminergic stimulant (exposure models), or a combination of both factors.

## Data Availability

No

## Acknowledgments

The authors acknowledge the Federación de Organizaciones no Gubernamentales de la Argentina para la Prevención y el Tratamiento de Abuso de Drogas (FONGA) as well as the patients, clinicians and operators of the therapeutic communities (Buen Sanmaritano, El Reparo, Foundation Creer es crear, Creando la libertad, El Palomar, Modelo Minnesota). Also, we acknowledge the incredibly generous and committed help of the people of the non -profit organizations who connected us with the CTR (Center Juan Pablo II, Foundation Temas, Matanza secretary, Agustin at Barrio Mitre).

